# Recurrent *ERBB2* alterations are associated with esophageal adenocarcinoma brain metastases

**DOI:** 10.1101/2025.02.19.25322558

**Authors:** Nora M. Lawson, Lingqun Ye, Chae Yun Cho, Bo Zhao, Thomas Mitchell, Inés Martín-Barrio, Bruno Beernaert, Archit Gupta, Matei Banu, Yonathan Lissanu, Sydney Shaffer, Hussein Tawbi, Jing Li, Maria Kristine Gule-Monroe, Christopher A. Alvarez-Breckenridge, Jason T. Huse, Mariella Blum Murphy, Feng Yin, Frederick F. Lang, Eileen E. Parkes, Jeffrey S. Weinberg, Kadir C. Akdemir

**Affiliations:** Department of Neurosurgery, MD Anderson Cancer Center, Houston, TX, USA; Department of Genetics, MD Anderson Cancer Center, Houston, TX, USA; Department of Oncology, University of Oxford, Oxford, UK; Department of Neurosurgery, Stanford University, Palo Alto, CA, USA; Department of Thoracic Surgery, MD Anderson Cancer Center, Houston, TX, USA; Perelman School of Medicine, University of Pennsylvania, Philadelphia, PA, USA; Department of Melanoma Medical Oncology, MD Anderson Cancer Center, Houston, TX, USA; Department of Radiation Oncology, MD Anderson Cancer Center, Houston, TX, USA; Neuroradiology Department, MD Anderson Cancer Center, Houston, TX, USA; Department of Gastrointestinal Medical Oncology, MD Anderson Cancer Center, Houston, TX, USA; Department of Anatomical Pathology, MD Anderson Cancer Center, Houston, TX, USA; Institute for Data Science of Oncology, MD Anderson Cancer Center, Houston, TX, USA; Department of Genomic Medicine, MD Anderson Cancer Center, Houston, TX, USA

**Author notes:** Co-first author (Nora M. Lawson, Lingqun Ye, Chae Yun Cho, Bo Zhao). Co-senior authors (Jeffrey S. Weinberg, Kadir C. Akdemir).

**Keywords:** Esophageal adenocarcinoma, brain metastasis, *ERBB2*, *JAK2*, micronuclei, ecDNA, tumor microenvironment, T cell infiltration, ADC therapies, immunotherapy

## Abstract

Brain metastases in esophageal adenocarcinoma (EAC) patients are associated with poor prognosis and remain understudied. We performed multi-omics analysis with whole-genome sequencing and single-cell spatial transcriptomics on the brain metastases and matched primary tumors. Our analysis identified *ERBB2* as a recurrent oncogene in EAC brain metastases, with 9 out of 10 cases harboring amplifications. Single-cell whole-genome and multi-region sequencing revealed that *ERBB2* alterations, occur early during disease progression and are associated with monoclonal seeding. Although the median survival in our cohort was 13 months, one patient on HER2 antibody-drug conjugate therapy remains a long-term survivor beyond 34 months. Interestingly, the sole patient without an *ERBB2* alteration had *JAK2* deletion, high T cell infiltration in the brain lesion, and survived 35 months after immune checkpoint therapy. Our findings have significant clinical implications for the treatment and management of EAC brain metastases.

**Highlights:**

- *ERBB2* is an early recurrent and targetable oncogene alteration in EAC-BM
- High T cell infiltration in *JAK2*-deleted tumor links to immunotherapy response
- Genomic instability of EAC-BM is marked by presence of micronuclei and ecDNA
- EAC brain metastasis resembles monoclonal seeding events

**Graphical Abstract:** 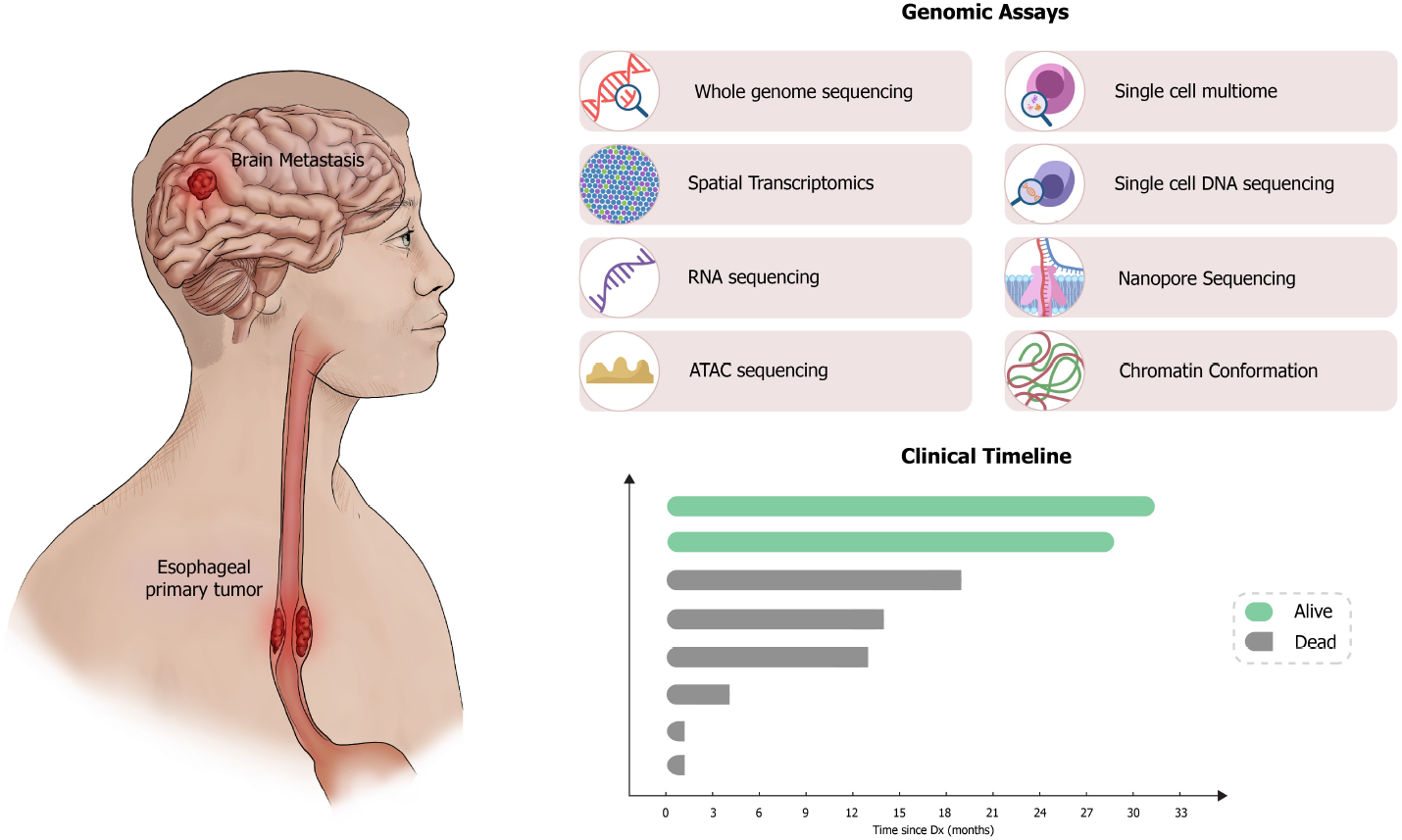

## Introduction

Esophageal adenocarcinoma (EAC) is a prevalent and fatal disease, ranking sixth globally for cancer-related deaths, with a 5-year survival rate of 15% and a rising incidence in the United States.^1^ EAC primarily affects white males, particularly those with gastroesophageal reflux disease or Barrett’s esophagus.^2,3^ While not all cases of Barrett’s esophagus progress to EAC, a progression pattern exists where the esophageal squamous epithelium can advance from non-dysplastic Barrett’s esophagus to low-grade dysplasia, high-grade dysplasia, and ultimately to EAC.^2^ Given this clinically well-characterized pre-cancer to adenocarcinoma progression, extensive research has been conducted on mutations and oncogene amplifications during the initiation of primary esophageal tumors from Barrett’s esophagus.^2,4-7^ One of the earliest driver events during this transition is the inactivation of the *TP53* tumor suppressor gene.^6^ The frequent mutations and inactivation of *TP53* gene in Barrett’s esophagus patients result in high tumor genomic instability, leading to high copy number alterations, whole-genome duplications, and single-nucleotide substitution rates.^6,8^ Moreover, frequent oncogene amplifications have been reported through the gain of chromosomal copies or extra-chromosomal DNA, resulting in high oncogene expression levels.^9, 10^ Esophageal adenocarcinoma commonly metastasizes to different sites including lymph nodes, liver, and lung; however, our understanding of molecular evolution of EAC metastases remains unclear.^11^ So far, the only genomic study of EAC metastases used bulk, multi-region whole-genome sequencing to evaluate the clonal organization of EAC metastases and suggested that EAC tumor metastases arise from multi-clonal seeding to various organs.^12^ However, a comprehensive evaluation of organ specific EAC metastases is currently not available. This is particularly true for brain metastases, where the infrequent occurrence of brain metastases in patients with EAC (2-6%) makes it challenging to study this tumor-type.^11, 13, 14^ In this study, we analyzed 10 EAC brain metastasis genomes, including seven with matched primary tumors, integrating multi-region and single-cell whole-genome data with spatial transcriptomics. This revealed recurrent *ERBB2* amplifications as a key oncogenic event, along with critical insights into clonal evolution and immune microenvironment changes from primary to brain metastasis. Our findings offer actionable therapeutic insights for this aggressive tumor subtype.

## Results

### *ERBB2* is a recurrent oncogene alteration in EAC brain metastases

To identify potential predictive biomarkers that could indicate a patient’s susceptibility to the development of brain metastases and to evaluate the changes in the tumor microenvironment during evolution of esophageal brain metastases, we analyzed 10 EAC brain metastasis genomes. Of these, eight underwent multi-omic profiling with whole-genome sequencing and spatial single-cell transcriptomics, while two published genomes^12^ were included for multi-region whole-genome sequencing analysis (Supplemental Table 1). The median survival after brain metastasis diagnosis was only 13.5 months in our cohort, consistent with the known poor prognosis of EAC patients with brain metastases.^11, 13, 14^ However, two patients in our cohort survived long-term, and are still alive 34- and 35-months following brain metastasis diagnosis, supporting the importance of understanding molecular underpinning of this disease for personalized treatment improving clinical outcomes for these patients.

To investigate the molecular features of EAC brain metastases, we compared the WGS data of 10 EAC brain metastases with that of the publicly available unpaired primary EAC International Cancer Genome Consortium Pan-Cancer Analysis of Whole Genomes (ICGC-PCAWG) datasets (n=85).^15^ We also compared our WGS data with that of the publicly available Hartwig Medical Foundation dataset comprising unpaired non-brain EAC metastases (n=140) most commonly to the liver (n=67) and lymph node (n=20).^16^ For our comparative analysis we primarily utilized results from a large-scale cancer genome reanalysis^17^ where a computational analysis pipeline is applied on samples from both the PCAWG and Hartwig datasets. We performed our analysis in a similar manner to this study so as to ensure that our results were comparable.

Our EAC brain metastases samples showed comparable single-nucleotide variation (SNV) and structural variation (SV) load compared with the primary tumor and metastases from outside the brain (Figure 1A). We assessed COSMIC single base substitution (SBS) mutational signatures present in the ICGC PCAWG primary EAC cohort, Hartwig EAC non-brain metastases cohort, and our brain metastasis cohort. The primary EAC cohort and EAC metastasis cohort exhibit signature SBS17a/b which has previously been reported as the predominant signature in this cancer type, characterized by T>G substitutions (Figure 1A).^18,19^Additionally, clock-like signatures (SBS1,5), proliferative signatures (SBS8, 18), and APOBEC signatures (SBS 2,13) were identified in each cohort (Figure S1A). We observed similar signature prevalence in EAC brain metastasis genomes with SBS17a/b as the predominant signature (Figure 1A).

**Figure 1:**
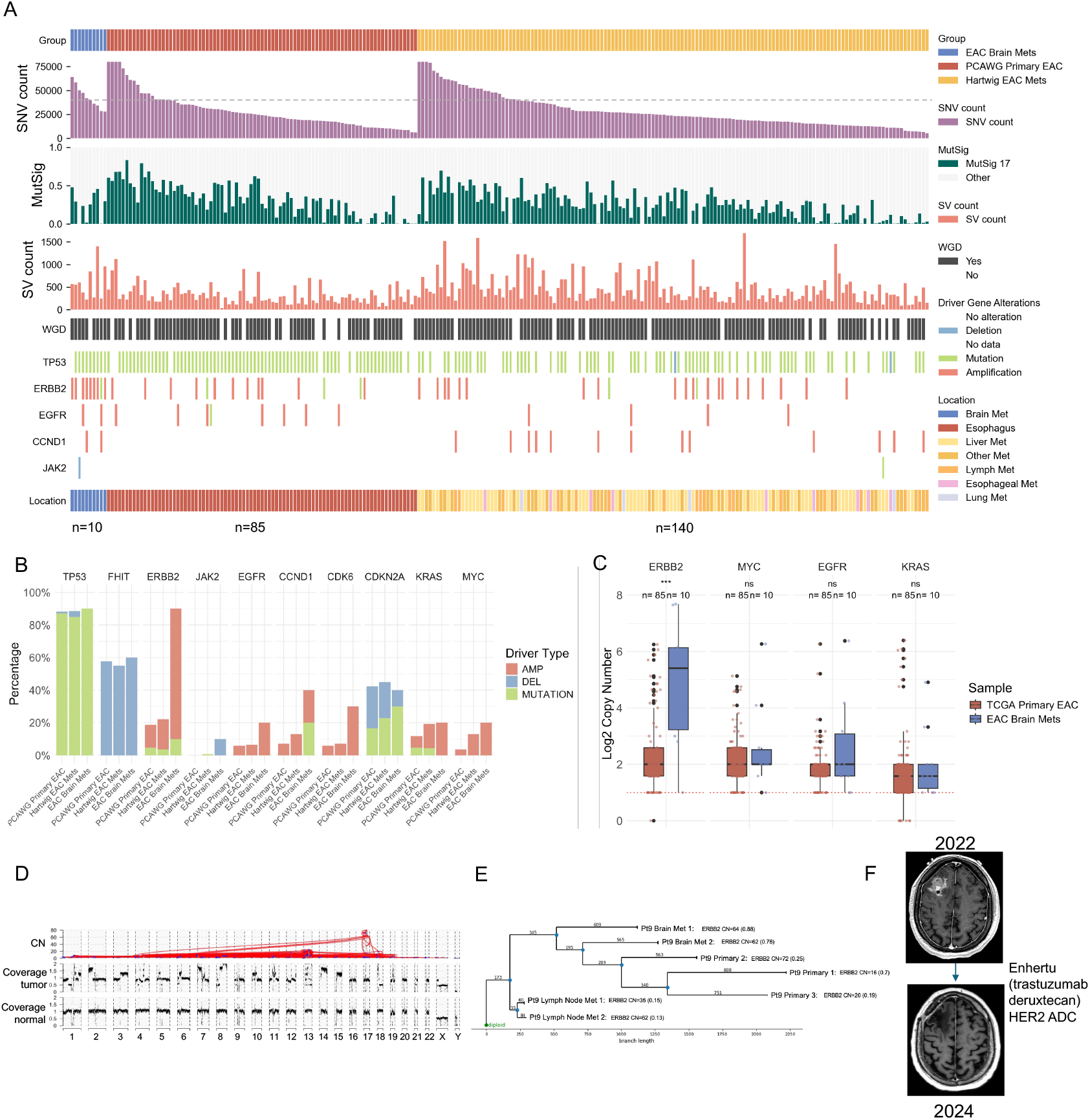
*ERBB2* is a recurrent oncogene alteration in EAC brain metastases, (A) Whole genome sequencing results comparing EAC brain metastasis cohort (n=10), PCAWG Primary EAC cohort (n=85) and Hartwig EAC metastasis cohort (n=140). Including single-nucleotide (SNV) count (gray line: y = 40,000), COSMIC mutational signature 17 contribution, structural variation (SV) count, whole-genome duplication (WGD) status, driver gene alteration, and location of sample. (B) Percentage of samples with a given oncogene indicated by type of oncogenic event comparing the three different cohorts (C) Log2 copy number for *ERBB2, MYC, EGFR*, and *KRAS* comparing TCGA primary EAC and EAC brain metastasis using a Wilcoxon rank sum test (*: p-value less than 0.05) (D) A representative EAC brain metastasis plot showing rearrangement, copy number (CN) and genome coverage (E) Phylogenetic analysis of multi-region WGS data from a patient (Patient 9) showing early alterations in *ERBB2* gene. Locations of biopsies included with copy number for *ERBB2* and purity of tumor sample. (F) MRI images of pre and post brain metastasis resection of Patient 2 who received HER2 ADC treatment (trastuzumab deruxtecan) See also Figure S1 and Table S1

The most recurrent driver mutation among these cases is *TP53* inactivation, suggesting this as the primary tumor-initiating event (Figure 1B).^2^ As a result, whole-genome duplication (WGD) is ubiquitous in the primary and metastatic EAC cases, including the brain metastases (Figure 1A).^2^ In addition, *FHIT* deletion has been reported as an early event during esophageal adenocarcinoma progression, and our data demonstrated that the ratio of *FHIT* deletion is similar in our brain metastasis cohort compared with the primary EAC cohort.^7, 19, 20^ Comparable findings were observed for other driver genes including *CDKN2A, MYC*, and *KRAS* (Figure 1B).^6,19^ Notably, *ERBB2* amplification driver gene occurrence is observed in 90% of EAC brain metastases. This percentage is markedly higher than in primary EAC and non-brain EAC metastases where only in approximately 20% is *ERBB2* indicated as a driver gene (Figure 1B).^6,19^ Next, we assessed the copy number levels of these select oncogenes and observed that *ERBB2* had a significantly higher (p-value < 0.0004) copy number in EAC brain metastases (CN range of 2 to 205) compared to TCGA primary EAC samples (CN range of 2 to 67) while other commonly amplified oncogenes had similar copy numbers in the two groups (Figure 1C). We also investigated whether the expression level of *ERBB2* was significantly higher in the EAC brain metastasis cohort compared with that of normal brain or esophagus tissues.^21^ We found that *ERBB2* has an expression level significantly higher (p-value < 0.001) in EAC brain metastases compared with primary EAC samples, normal esophagus, and normal brain tissues (Figure S1B).

To investigate whether *ERBB2* amplifications occur in primary EACs we generated targeted oncogene sequencing from matched primary tumors (n=5). From targeted sequencing data, we determined that *ERBB2* amplifications were present in all available matched primary tumors, indicating that *ERBB2* amplifications occur early in tumor evolution before brain metastases arise (Figures S1C). We further investigated the evolution of *ERBB2* in two patients using multi-region WGS data^12^ including data from primary EACs, lymph nodes, and EAC brain metastases. We detected *ERBB2* amplifications present in all the samples from these two patients, and associated large structural rearrangements were identified in chromosome 17 (Figure 1D, S1D). Reconstructing the phylogenies for these two patients via copy number variations,^22^ we found that for both patients the brain metastasis samples were closely related to the primary samples indicating that *ERBB2* amplifications are early events that occur before the tumor metastasizes to the brain, as high *ERBB2* amplifications were shared with all the samples (Figure 1E, S1E).

These results are particularly significant given that development of novel antibody-drug conjugates targeting *ERBB2* are approved for patients with brain metastases. Most notable is trastuzumab deruxtecan which has shown clinical benefit in patients with brain metastasis from breast cancer.^23^Although some of our patients were treated with HER2 targeted therapies, particularly trastuzumab (which does not cross the blood brain barrier), only one patient (Patient 2) in our cohort was treated with trastuzumab deruxtecan. This patient experienced a deep and durable response, receiving treatment for 22 months, and is one of only two long-term survivors in our cohort (Figure 1F, Supplemental Table 1). This case is particularly notable since this patient had leptomeningeal disease,^24^ which is associated with an extremely poor outcome warranting whole-brain radiation. Our identification of early and recurrent *ERBB2* alterations in EAC brain metastases, along with a case of successful treatment using a HER2 antibody-drug conjugate, underscores the critical clinical implications for the screening and treatment of patients with EAC brain metastases.

### High T cell infiltration in brain metastasis with *JAK2* deletion successfully treated with immunotherapy

Tumor microenvironment is an important component of disease progression.^25^ To investigate changes in the tumor microenvironment of EAC brain metastases and explore potential crosstalk with cancer cell genomic drivers, we employed a single-cell spatial transcriptomics approach (10x Genomics Xenium). This included two unique pre-designed panels—the Xenium human brain panel and the Xenium human immune-oncology panel—plus a custom 100-gene panel added to the human brain panel (Figure 2A, Supplementary Table 1). We profiled 8 brain metastases, 5 matched primary tumors, and 2 metastases from the lymph node and liver. In total, 746 genes were analyzed across 1,729,039 cells, yielding over 265 million transcripts. (Figure 2A). Cell types within the microenvironment were determined by marker gene expression patterns with identification of malignant cells, neuronal cells, various types of immune cells, and vascular cells based on the human brain and custom gene panel (Figure S2A, B, Supplementary Table 2).

**Figure 2:**
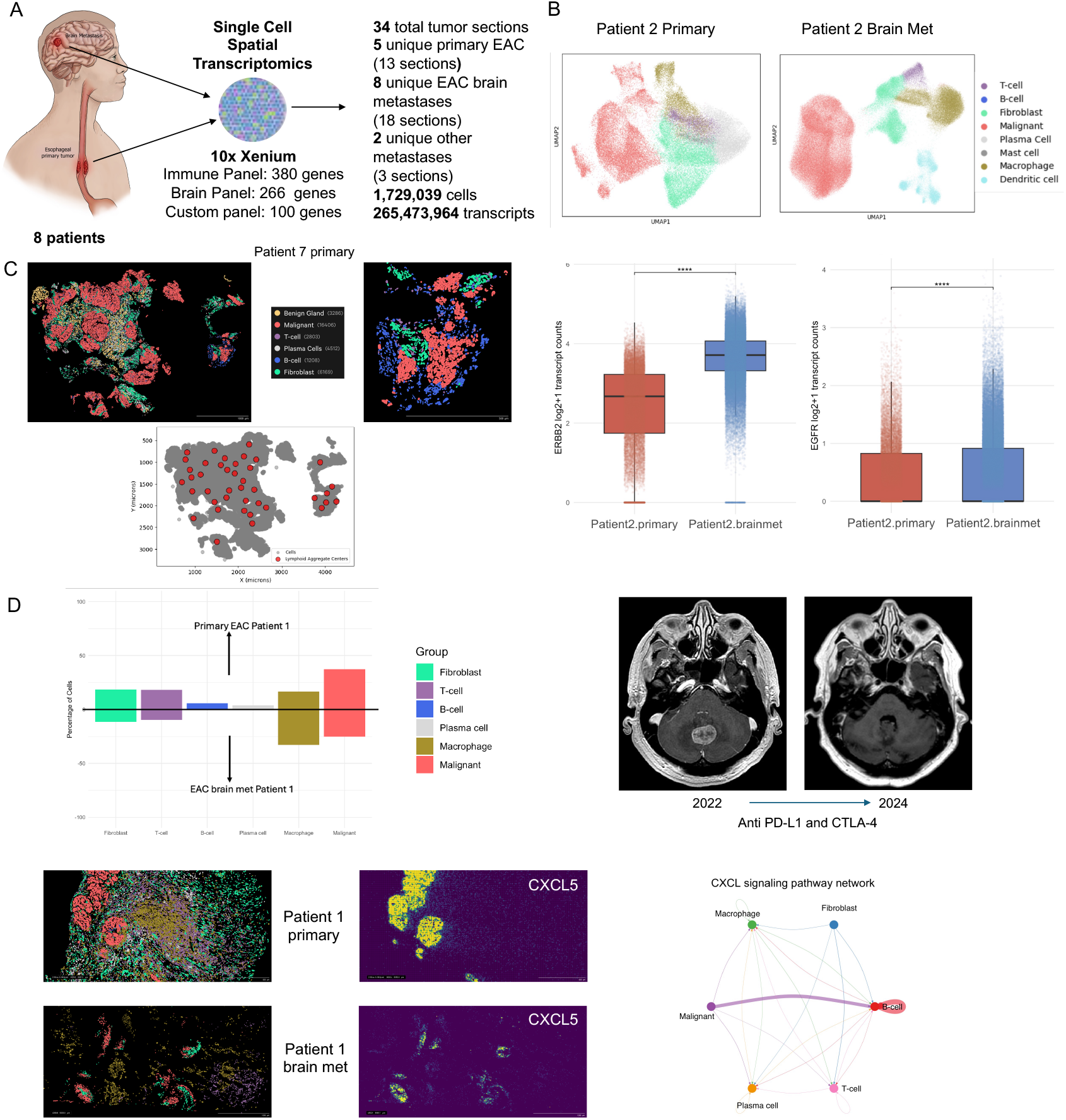
Immune cell infiltration in primary tumors and high T cell infiltration in brain metastasis with *JAK2* deletion treated with immunotherapy, (A) Graphical summary of spatial single-cell transcriptomics results (B) (upper panel) Patient 2 primary tumor and brain metastasis spatial organization is shown with UMAP (lower panel) *ERBB2* and *EGFR* expression count comparison between the primary and brain metastasis tumor sections in malignant cells (C) Patient 7 primary tumor spatial maps display the primitive tertiary lymphoid structures/lymphoid aggregates (as red dots in below plot) (D) Bar plot shows cell type proportion comparison between primary and brain metastasis. MRI images pre and post brain metastasis resection of Patient 1 who has been receiving anti PD-L1 and CTLA-4 therapies. Patient 1 primary tumor and brain metastasis spatial organization is shown with example spatial plots. (bottom right) Patient 1 primary tumor and brain metastasis spatial maps show density of expression for *CXCL5* and CellChat circle plot displays CXCL signaling network for Patient 1 primary. See also Figure S2 and Tables S2, S3

Our immune oncology panel analysis allowed us to identify T cells (∼1.6%), macrophages (∼12%), and dendritic cells (∼2%) in brain metastasis tumors, and T cells (∼11.1%), B cells (∼2.9%), macrophages (∼10.5%), plasma cells (∼11.6%), and mast cells (∼0.8%) in matched primary tumors (Figure S2B, Supplementary Table 2). Our initial comparative analysis indicated that primary EAC tumors have a higher immune cell population (∼36.9%) than matched brain metastases (∼16%) (Figure 2B, S2B, Supplementary Table 2).^26^ Expectedly, we detected more neuronal cells (∼12.6%) in brain metastasis compared with matched primary tumors (0%) (Supplemental Table 2). This result is also confirmed in our bulk RNA-seq comparison between EAC brain metastases with primary EAC tumors (Figure S2C).

We also investigated oncogene expression levels between matched primary and brain metastasis. We observed that *ERBB2* expression was significantly higher (p-value < 0.01) in the brain metastases malignant cells in Patient 2 (median value is 12.1 compared to primary tumor median value of 5.4). Whereas *EGFR* expression levels in malignant cells were comparable (p-value < 0.01) across primary tumor and brain metastases in Patient 2 (median value is less than 1 for both primary and brain metastases sample) (Figure 2B).

Higher lymphoid structures comprised of T cells and B cells, are associated with response to anti-tumor therapies.^27^ In our samples, we observed lymphoid aggregate organization of T cells and B cells in primary tumors, however such organization in brain metastases was rarely observed. Within the primary tumor from Patient 7 we observed primitive formations of tertiary lymphoid structures (TLS) utilizing a recent algorithm,^28^ in which B cells and T cells formed a lymphoid aggregate juxtaposed to malignant cells (Figure 2C, S2D). In contrast, the matched brain metastasis of Patient 7 did not contain significant immune infiltrate. This patient was not treated with any immune-checkpoint blockage therapies and passed away within 2 months of the diagnosis of brain metastasis. On the other hand, the primary tumor of Patient 1 also had lymphoid aggregates and this patient’s brain metastasis also had the highest level of T cell infiltration in our cohort (∼10%) which was obtained pre-immunotherapy treatment (Figure 2D, S2E). Consistent with this, expression of the TLS marker *CXCL13* was elevated within T cells from this tumor (Figure S2F). Interestingly, this patient was treated with anti-PD-L1 and anti-CTLA4 immunotherapies and is now still alive 35 months after the diagnosis of brain metastasis (Figure 2D).

The unique tumor microenvironment in this sample prompted us to further investigate the driver events underlying this patient’s tumor. Notably, this tumor was the only non-*ERBB2* amplified cancer in our cohort. Instead, we detected a *JAK2* deletion as one of the main oncogenic events, along with a *CDK6* amplification (Supplementary Table 3). This tumor also had none of the signature 17 mutational profile which is common to EAC tumors (Figure 1A).^18^ However, the histopathological analysis confirmed this tumor as EAC. Given the successful immunotherapy response rate in this patient, we further investigated *JAK2* alterations in primary and other EAC metastatic datasets. This alteration is rare as we could detect only a single EAC metastasis to liver with *JAK2* driver mutation in the Hartwig cohort. Additionally, we investigated *CD274*, which codes for PDL1 and is located near *JAK2*, to determine whether this gene was also affected by the deletion on chromosome 9. While Patient 1 exhibited the lowest expression of *JAK2*, PDL1 expression was above average compared to other samples in the cohort, suggesting that PDL1 was likely not affected. (Figure S2G). Interestingly, previous reports have suggested *JAK2* mutations as a mechanism of immunotherapy resistance in melanomas.^29,30^ However, more recently JAK pathway activation has been identified as a resistance mechanism to immune checkpoint therapy in solid tumors with re-sensitization to immune checkpoint blockade achieved with therapeutic JAK inhibition.^31, 32^

We additionally conducted ligand-receptor analysis^33^ utilizing the single cell resolution spatial data to investigate interactions between cytokine ligand and receptor pairs in primary EAC tumors with larger immune populations. Specifically, we aimed to identify differences between Patient 1, which had the largest immune cell population (44.2% of cells), with other primary EAC tumors. CCL and CXCL signaling pathways emerged as significant in all three tumors analyzed (Figure 2D, S2H-K). Our analysis revealed robust cell-to-cell communication in tumors with lymphoid aggregates, particularly CXCL signaling, where malignant cells primarily sent signals to B cells (Figure 2D, S2H-K). CCL signaling instead primarily involved interactions amongst immune cells, including T and B cells in addition to T cell autocrine signaling (Figure S2H-K).

In Patient 1, we observed increased interactions between T cells and macrophages in the CCL pathway, while in other samples, T cell and B cell interactions dominated (Figure S2H). Notably, Patient 1 exhibited higher overall expression of ligand-receptor pairs in both CXCL and CCL pathways (Figure 2D, S2H, I). Specifically, T cells showed elevated expression in the CCL pathway, while malignant cells and B cells showed heightened activity in the CXCL pathway (Figure 2D, S2H, I) Additionally, Patient 1’s sample demonstrated increased contribution of the *CXCL5-CXCR2* pair, which was absent in the other cases (Figure 2D). In gastric cancers, increased expression of *CXCL5* has been shown to facilitate epithelial-to-mesenchymal transition (EMT), enhancing the metastatic potential of tumor cells.^34^ In this case, involving the only HER2-negative brain metastasis, these findings suggest that elevated cytokine expression in this tumor may have facilitated metastatic spread.

### Chromosomal instability of EAC brain metastasis tumors is marked by presence of micronuclei and ecDNA

Esophageal adenocarcinoma cases represent high levels of chromosomal instability^2^ as evidenced by the presence of micronuclei structures and extra-chromosomal DNA amplifications.^10^ The 10x Xenium spatial transcriptomics platform performs DAPI staining for detection of nuclear segments as a regular part of the analysis workflow. While analyzing our spatial single-cell transcriptomics data, we realized there were smaller DAPI signals within malignant cells, suggestive of micronuclei structures. Because the Xenium protocol involves ATP1A1/CD45/E-cadherin for cell boundary segmentation, and that the identified micronuclei-like structures are notably smaller compared with nuclear DAPI staining, and are only observed in malignant cells, we hypothesized that Xenium DAPI staining could be used to identify micronuclei in tumor sections (Figure 3A, S3A). To validate this observation, we performed DNA fluorescence in situ hybridization (FISH) against probes targeting the *ERBB2* region on an adjacent slide using a brain metastasis sample from Patient 7 (Figure 3A, S3B, C). This analysis confirmed the high *ERBB2* signal in smaller DAPI regions suggesting these regions contain high levels of oncogene amplifications. As an orthogonal validation, we performed cGAS and STING immunofluorescence staining on adjacent tissue sections from the same tumors.^35^ Given the small size of micronuclei (typically < 1 µm), adjacent tissue sections are unlikely to contain the same tissue structure. However, within the same regions of interest we identified high numbers of cGAS-positive micronuclei, consistent with high levels of chromosomal instability (Figure 3A, S3D). STING staining was notably absent in tumor regions and expression was restricted to stromal regions (Figure S3D). Taken together, we demonstrate sensitive identification of altered nuclear structures using Xenium staining that enables identification of regions of chromosomal instability using this approach. This analysis suggests that, in addition to spatial single-cell transcriptomic analysis, the imaging data generated through the Xenium workflow permits novel analyses such as micronuclei detection.

**Figure 3:**
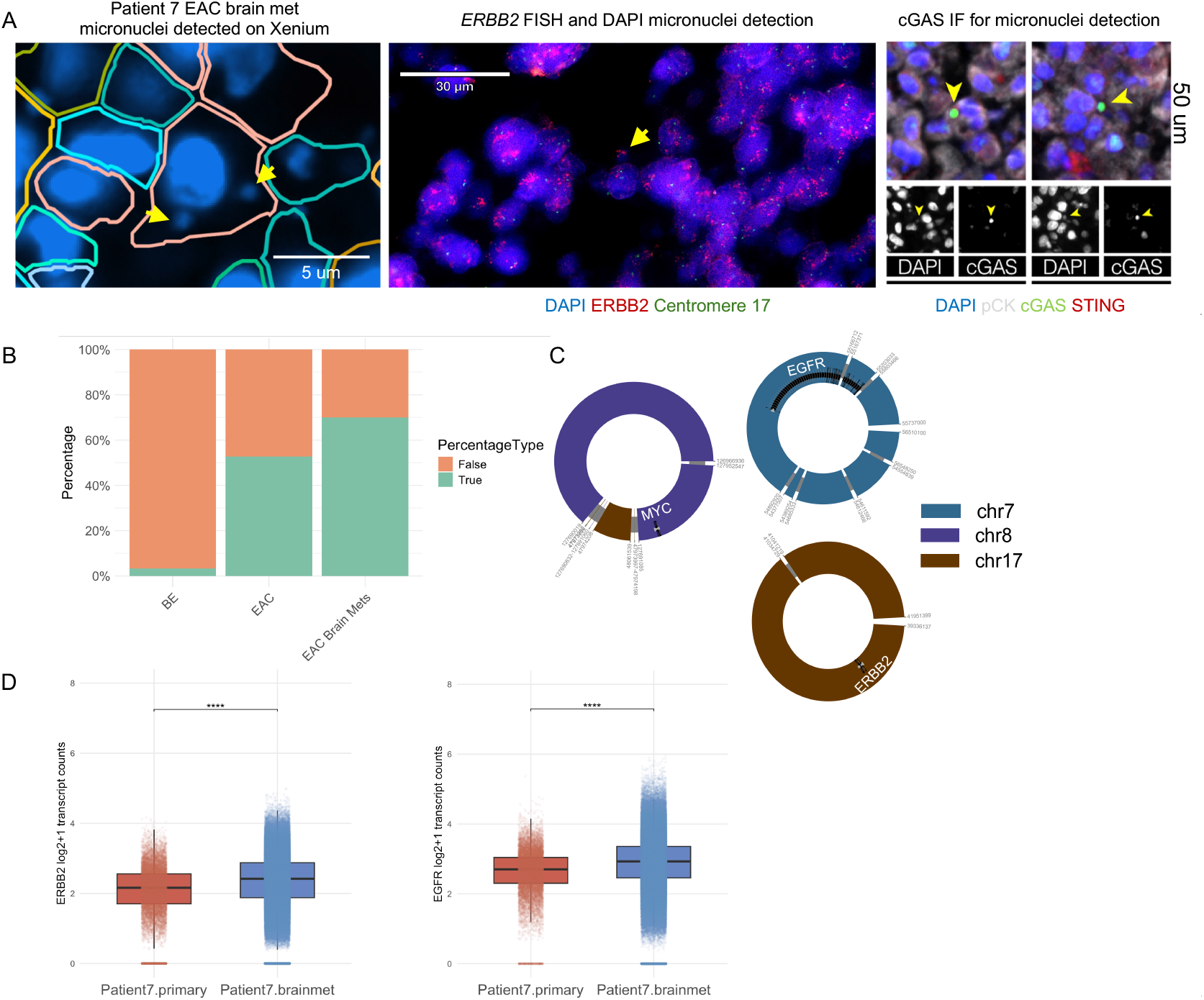
Chromosomal instability of EAC brain metastasis tumors is marked by presence of micronuclei and ecDNA, (A) Example micronuclei identified in Patient 7 from Xenium Explorer (10x Genomics), and additional micronuclei present in adjacent sections for Patient 7 confirmed with *ERBB2* fluorescence in situ hybridization (red) and cGAS immunofluorescence staining (green) (B) Bar plot shows percentage of samples with at least 1 ecDNA present in Barrett’s Esophagus (BE), primary EAC^10^ and EAC brain metastasis cohorts (C) Reconstruction of 3 separate ecDNA molecules detected in Patient 7 brain metastasis genome (D) Bar plots show Patient 7 Xenium single cell *ERBB2* and *EGFR* transcript count comparing primary and brain metastasis sample using a Wilcoxon rank sum test See also Figure S3

Another important characteristic of esophageal adenocarcinomas is high levels of extra-chromosomal amplifications (ecDNA). *ERBB2* is one of the main oncogenes observed in the ecDNA regions.^10^ To understand the nature of oncogene amplifications in our brain metastasis cohort, we applied Amplicon Architect and Amplicon Classifier algorithm^36^ for detecting ecDNAs. This revealed that 7 of 10 patients had at least one ecDNA molecule (Figure 3B). For *ERRB2* amplifications, we observed distinct types of alterations including ecDNA (n=1), linear amplification (n=4), and bridge-fusion-bridge amplification (n=4) in our main cohort (Figure S3E).

Interestingly, one brain metastasis sample (Patient 7) contained three distinct ecDNA species predicted by Amplicon Architect algorithm where each encompassed important oncogenes, namely *EGFR, ERBB2*, and *MYC* (Figure 3C, S3F). We also applied the chromatin capture technique (Hi-C) on the same tumor, revealing chromatin interaction patterns reported from ecDNA molecules (Figure S3G). To rule out that the detected unique ecDNA molecules were not due to lack of power secondary to short-read sequencing data, we generated high-coverage whole-genome sequencing data from this tumor using the Oxford Nanopore long-read sequencing technology. Applying a recent ecDNA reconstruction algorithm^37^ validated that the observed ecDNA molecules were not overlapping and indeed were separate molecules (Figure S3F).

To understand the clonal organization of these amplifications, i.e., whether they reside on different cells or within the same cell, we first investigated the single-cell spatial transcriptomic data from this tumor. Notably, for Patient 7, the spatial transcriptomic data and the scRNA sequencing data revealed cells co-expressing *ERBB2* and *EGFR* in the brain metastasis (Figure 3D, S3H). These data support a recent work suggesting distinct ecDNA molecules can co-segregate during cell division.^38^ This comparison of cells expressing *ERBB2* and *EGFR* genes differs when analyzing tumors lacking *ERBB2* and *EGFR* ecDNAs (for example Patient 2). In these tumors, we observed a significant difference in transcription levels between *ERBB2* and *EGFR* (Figure 2B). However, in Patient 7 we observed that distinct ecDNA species exhibit similar transcription profiles (Figure 3D). Our analysis demonstrates that the previously proposed ecDNA co-segregation phenomenon in cancer cell lines can be partially recapitulated in tumor sections using a single-cell spatial transcriptomics approach. Our findings highlight the utility of this technology for further investigating ecDNA dynamics within the tissue context.

### Early and monoclonal metastatic seeding events

Given the unique case of three ecDNA species in a tumor, we sought to investigate the evolution of clones from primary EAC to brain metastasis. We performed single-cell whole-genome sequencing on samples from Patient 7. Our analysis identified 1418 cells that passed a quality control filter developed by an earlier study.^39^ Among these 1418 cells, 52 were from the primary tumor and 1366 were from the brain metastasis. The low level of detected primary cells was due to the FFPE tissue being the only available source of tissue for this primary tumor. This analysis revealed that all the malignant cells from the brain metastasis harbored the three unique ecDNA molecules, supporting our spatial transcriptomic findings (Figure 4A-C). We also performed single-cell multi-ome on the brain metastasis and copy number analysis^40^ based on single cell ATAC sequencing showed the co-occurrence of *EGFR, ERRB2*, and *c-MYC* region amplifications in the same cells (Figure S4A). These results suggest that distinct ecDNA molecules can co-segregate. Because the majority of cells harbor all three ecDNA molecules, these amplifications are likely driving co-selection, as suggested by a recent study.^38^ From the scWGS results we found that the malignant cells from this patient’s primary tumor also showed the same genomic abnormalities, where *EGFR* and *ERBB2* ecDNA amplicons were present. We did not detect the *MYC* ecDNA amplicon in the primary tumor, however, we cannot rule out if this was due to the low cell count isolated from the primary tumor tissue (Figure 4A). Copy number analysis of individual genes from scWGS of Patient 7 indicated that there was a correlation between high copy number of *EGFR* and *ERBB2*, confirming that the same cells contain both amplifications with the brain metastasis sample containing a higher copy number of *EGFR* and *ERBB2* when compared with the primary sample (Figure S4B). This data indicates multiple oncogenic changes exist in single cells, and the brain metastatic event originated from a mono-clonal mechanism.

**Figure 4:**
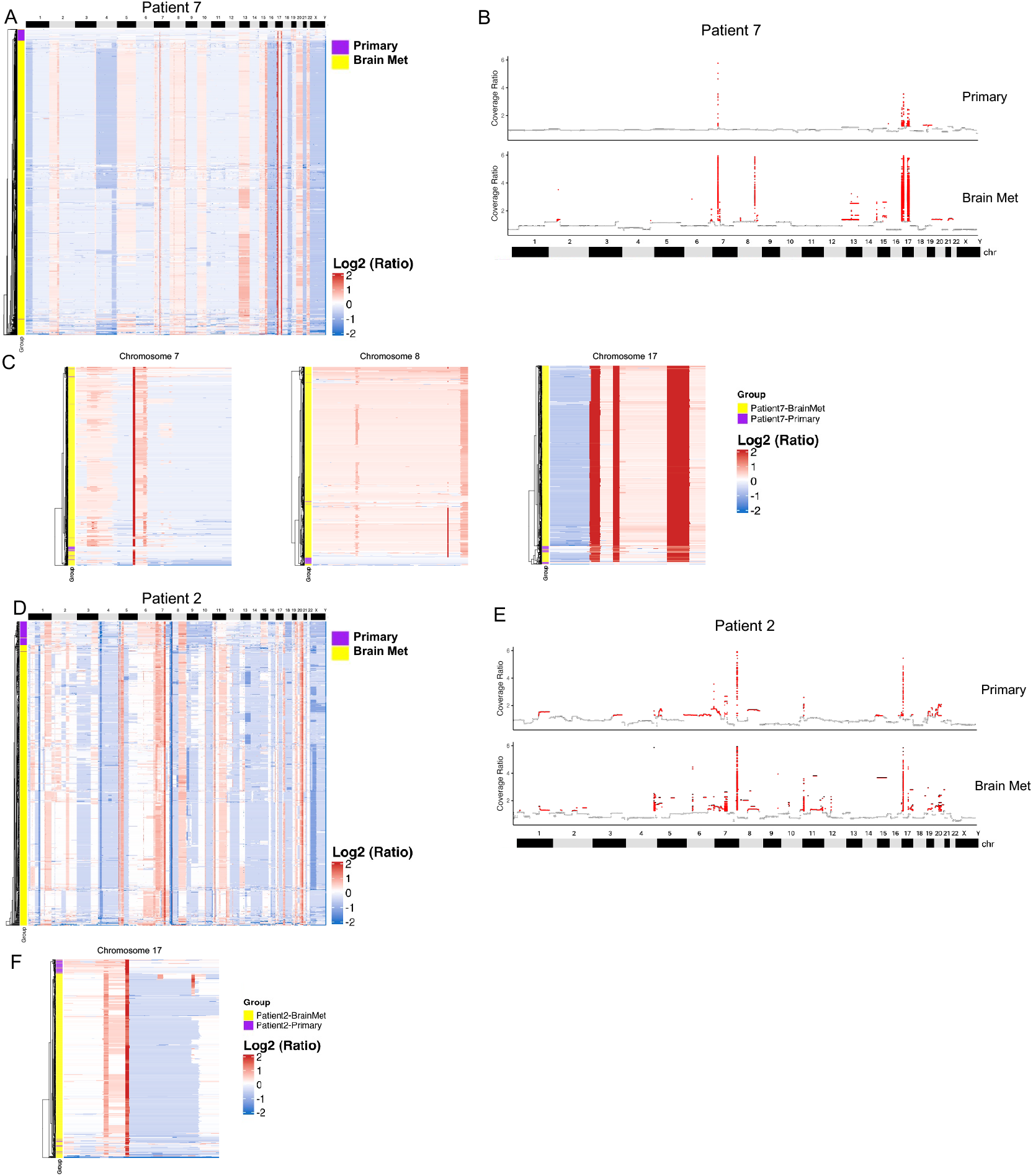
Early and monoclonal metastatic seeding events, (A) scWGS heatmap of log2 binned coverage ratio per chromosome for Patient 7 primary and brain metastasis samples. (B) Pseudobulk coverage ratio by genomic bins for Patient 7 primary and brain metastasis with amplified regions highlighted in red. (C) Heatmaps highlight amplification on chromosomes 7, 8 and 17 for Patient 7 (D) scWGS heatmap of log2 binned coverage ratio per chromosome for Patient 2 primary and brain metastasis samples. (E) Pseudobulk coverage ratio by genomic bins for Patient 7 primary and brain metastasis with amplified regions highlighted in red. (F) The heatmap highlights amplification on chromosome 17 for Patient 2 See also Figure S4

To expand our finding of early and monoclonal seeding of metastatic tumor cells in the brain, we additionally performed the same single-cell whole-genome sequencing profiling from the primary and metastatic tumors of Patient 2. This analysis showed similar evolutionary organization where the dominant clone in the brain metastasis was detected in the primary tumor, and the brain metastasis had further copy number alterations, suggesting an ongoing genomic instability and evolution (Figure 4D-F, S4E, F).

## Discussion

Our study elucidates critical molecular events in the development of brain metastases from esophageal adenocarcinoma, including the importance of *ERBB2* amplifications, likely early metastatic seeding, and the monoclonal nature of metastatic events. These findings have significant implications for targeted therapies and strategies to detect and manage brain metastasis in patients with EAC.

One of the most striking observations from our cohort is the prevalence of *ERBB2* amplifications in EAC brain metastases, identified in 90% of our cases.^41-44^ This rate far exceeds the 20% prevalence reported in primary EAC^6,19^, suggesting that *ERBB2* amplification is a critical driver event that may predispose EAC tumors to metastasize to the brain. Our findings also indicate that *ERBB2* amplification is associated with a significant increase in expression levels compared to normal brain and esophageal tissues, making it a promising therapeutic target. Importantly, *ERBB2*-targeted antibody-drug conjugates such as trastuzumab deruxtecan, which are known to have intracranial activity with clinical efficacy in other ERBB2-driven metastases, hold potential as a viable therapeutic option for EAC brain metastases.^23^ Notably, the only long-term survivor with an *ERBB2* amplification was treated with trastuzumab deruxtecan. This case underscores the potential of brain-penetrant *ERBB2*-targeted ADCs to improve outcomes for EAC patients with brain metastases, warranting further investigation into these agents’ efficacy in this context.

The phylogenetic analysis reveals that EAC brain metastases likely result from early metastatic seeding events, with *ERBB2* amplification present in primary tumor samples before metastasizing.^45^ This finding suggests that once *ERBB2* amplifies, these cells have an increased capacity to invade the brain, which complements findings from studies of breast cancer brain metastases.^46^ Such early seeding reinforces the importance of proactive brain screening in high-risk EAC patients, particularly those with *ERBB2*-amplified primary tumors.^47^ Early detection of brain metastases could enable timely interventions, potentially improving survival rates in these patients.

The single-cell genomic analyses reveal that brain metastases in our EAC cohort arise through monoclonal seeding, where a single clone from the primary tumor initiates metastatic spread. This clonal origin suggests that, despite ongoing genomic instability, a dominant metastatic clone is responsible for establishing and evolving brain metastases. These results highlight that brain metastases in EAC are likely driven by a select population of primary tumor cells that retain proliferative activity and harbor metastatic potential.^47^ This finding has clinical implications, emphasizing the importance of targeting specific clones to prevent and treat metastases. Single-cell multi-omic approaches provide insight into the clonality of metastatic EAC cells, enabling personalized strategies that target the defining mutations and copy number alterations characteristic of these dominant clones.

In one exceptional responder to immune checkpoint therapy, we identified deletion of *JAK2* as an event occurring in the primary tumor that was consistent in the brain metastasis. Although this appears counterintuitive, recent reports suggest that in a chronically inflamed tumor microenvironment, constant interferon stimulus results in terminally exhausted T cells with poor tumor control.^48^ Similarly, the development of EAC typically occurs in an inflamed background^2^, and in this setting JAK inactivation could permit paradoxical immune response and effective tumor control in response to immunotherapy, as observed in this patient. We only identified one other instance of this mutational event within an EAC liver metastases suggesting this as a rare occurrence. However, the striking response observed in this case suggests that further investigation of JAK-targeting in this typically inflammatory tumor could be warranted as a therapeutic strategy.

In summary, our study demonstrates *ERBB2* amplifications as a targetable driver in EAC brain metastases. It underscores the importance of early detection and intervention strategies for patients at high risk of metastatic spread to the brain. The monoclonal nature of these metastatic events highlights the potential of clonal targeting therapies, coupled with early screening and *ERBB2-*directed treatments, which could transform the therapeutic landscape for EAC brain metastases.

## Limitations of the study

The cohort for this study was small due to the rarity of this tumor subtype. However, we conducted a multi-omics analysis incorporating multiple data types per sample and included matched primaries when available. Additionally, we compared our findings to previously published large datasets to provide broader context and validation. Further studies with extended EAC brain metastasis cohorts will reveal additional findings.

The use of single-cell spatial data with probe-based panels posed challenges in cell annotation due to the limited number of probes. To improve tumor microenvironment profiling, we applied two distinct panels to adjacent tumor sections. This approach involves using multiple tissue sections from the same tumor block. While data integration is possible, future technological advancements will enable the profiling of a higher number of genes within the same tissue sections.

## Data Availability

The raw bulk data reported in this study will be deposited in dbGaP. Any additional information is available from the lead contact upon request.

## Resource Availability

### Lead Contact

Further information and requests for resources should be directed to and will be fulfilled by the lead contact, Kadir C. Akdemir.

### Material Availability

This study did not generate new unique reagents.

### Data and Code Availability

The raw bulk data reported in this study will be deposited in dbGaP. Analysis scripts are deposited in the Akdemir Lab GitHub: https://github.com/akdemirlab/EAC-brainMets. This paper does not report any original code. Any additional information is available from the lead contact upon request.

## Acknowledgements

We thank the patients and their families for contributing to this study. We thank Linghua Wang and Peter Van Loo for their critical reading of this manuscript. We would also like to thank the Brain Tumor Center pathology department, especially Lisa Norberg, Truc Kuo, and Jennifer Ritchie, for their essential contributions to patient sample collection and preparation. This work was supported by Andrew M. McDougall Brain Metastasis Clinic and Research Program Award (K.C.A), SPORE in Brain Cancer CEP (P50CA127001 to K.C.A.), Kleberg Innovative Investigator Award (K.C.A), Sachs Family Funds (J.S.W).

## Author contributions

Formal Analysis, N.M.L.; Data Curation, N.M.L. and L.Y.; Software, N.M.L., L.Y., B.Z., A.G.; Investigation, I.M.-B., C.Y.C., B.Z., B.B., and T.M.; Visualization; N.M.L., I.M.-B., B.Z., L.Y., T.M., B.B.; Writing – Original Draft, N.M.L. and K.C.A.; Writing – Review & Editing, N.M.L., K.C.A., J.S.W., E.E.P., and Y.L.; Conceptualization, K.C.A. and J.S.W.; Methodology, K.C.A; Supervision, K.C.A., J.S.W., Y.L., S.S., H.T., J.L., M.K.G.-M., C.A.A.-B., J.T.H., M.B.M., F.Y., F.F.L., E.E.P.; Resources, J.S.W., M.B., J.T.H., F.Y.; Project Administration, K.C.A., Funding Acquisition, K.C.A.

## Supplemental information

**Document S1**. Figures S1–S4, related to Figures 1-4

## Supplemental Tables.xlsx

Table S1. Summary of data types and clinical data available for each patient, related to Figure 1

Table S2. Xenium cell type ratios per patient sample for brain and immune panels, related to Figure 2

Table S3. Patient 1 Brain Metastasis Driver Genes, related to Figure 2

## Methodology

### Study Design

We performed a thorough prospective evaluation of EAC brain metastasis resections and frozen specimens collected through the CATALYST pipeline initiated by the Department of Neurosurgery. All patients provided written informed consent under protocols approved by the Institutional Review Board of MD Anderson, in accordance with ethical standards for genomic research. The pathological evaluation confirmed that these tissues were brain metastases from primary esophageal adenocarcinomas. DNA and RNA extraction was performed using standard methods for FF and FFPE tissues. Illumina whole genome sequencing (WGS) (tumor: 60x, normal: 30x), bulk RNA-sequencing, and Agilent cancer panel sequencing (coverage 500x) was generated by UT MD Anderson Moonshot Program Cancer Genomics Laboratory using standard Illumina library preparations and sequenced on Illumina NovaSeq platform. Oxford Nanopore Technology (ONT) long read sequencing was performed on the PromethION platform by MD Anderson Advanced Genome Sequencing Core. Single nuclei DNA sequencing for 2 pairs of primary and brain metastasis samples was performed by the CPRIT single cell genomic center (SCGC) at MD Anderson Cancer Center. 10x Genomics Xenium spatial transcriptomics were performed with our in-house machine as described below.

### Whole-genome sequencing Analysis

We used BWA-mem (v0.717) algorithm^49^ to map sequencing reads against the GRCh38 reference genome. We sorted, indexed, and marked duplicates in BAM files using Picard (v3.1.0)^50^ and SAMtools (v1.15)^51^. Tumor ploidy and purity was determined using ASCAT (v3.1.2)^52^ and PURPLE (HMF tools v3.9)^53^ software. For multi-region sequencing comparisons, we used Refphase (v0.20)^54^ and MEDICC2 (Minimum Event Distance for Intra-tumour Copy-number Comparisons)(v0.9.2)^22, 55^ to determine somatic copy number and develop copy number phylogenies to better understand the EAC genome evolution.

We compared our results with publicly available data on genomic characterizations of primary esophageal adenocarcinoma tumors. WGS results from Hartwig Medical Foundation dataset^16^, PCAWG datasets^15^, and TCGA datasets^56^ were included in the analysis of a previous publication^17^. We performed our analysis the same as the study so that we could ensure that our results were comparable using LINX pipeline (HMF tools v3.9)^16, 53^ for both SNV and SV calls. SNVs were called using SAGE somatic (v2.2; HMF tools v3.9), and the SVs were called using PURPLE. Driver genes were also determined using PURPLE with the driver genes filtered for greater than 0.5 likelihood. PURPLE’s whole genome doubling prediction was used to compare the percentage of samples with WGD to the same groups. To compare the driver gene copy number levels between our cohort and the TCGA primary EAC cohort, we used ASCAT (v3.1.2)^52^ segmentation copy number data and applied a Wilcoxon rank sum statistical test. We also used JaBbA software (v1.1)^57^ for visualizing copy number and structural variation information for each tumor. We performed single-nucleotide signature analysis by utilizing EnsembleFit (Consensus SBS Mutational Signature Assignment).^58^

### ecDNA Detection

To classify oncogene amplifications, we used AmpliconSuite-pipeline (v1.0.0)^36^. Copy number variations were identified using CNVKit^59^ and used as an input for AmpliconArchitect algorithm.

AmpliconClassifier function was used to determine the type of focal amplifications that occurred because of ecDNA, BFB, linear amplification or none. We calculated the percentage of samples with at least 1 ecDNA present within our EAC brain metastasis samples and compared these percentages with two other cohorts including Barrett’s esophagus and esophageal adenocarcinoma cohorts utilizing results from Luebeck et al. 2023^10^ study.

### Nanopore Sequencing Analysis

Oxford Nanopore long read sequencing reads were aligned with GRCh38 using Minimap2 (v2.28).^60^ We used SAMtools^51^ for sorting, adding MD tags with calmd, and indexing the aligned bam file. To compare our short-read based ecDNA detection, we used Decoil pipeline (v1.1.2-slim)^37^ to reconstruct ecDNA using long-read data. The decoil-pipeline performs SV calling, coverage track, and reconstruction. This pipeline was paired with decoil-viz (v1.0.3) which is utilized for visualization of the ecDNA amplicons.

### Bulk RNA Analysis

We used STAR (v2.7.9a)^61^ algorithm for mapping the reads to the GRCh38 reference genome, SAMtools (v1.15)^51^ for filtering the reads, and Deeptools (v.3.1.3)^62^ for bam file coverage. The result was a gene output tab file and a bam file which can be used for downstream analysis. DeSeq2 (v.3.18)^63^ is a Bioconductor R package that we used to determine the TPM (transcripts per million) and normalize the data using the reads per gene output tab file from STAR. This analysis normalizes the expression data so that further analysis can be performed. We also performed pathway enrichment analysis using Gene Set Enrichment Analysis (GSEA).^64, 65^ The Genotype-Tissue Expression (GTEx) Portal^21^ was employed to obtain *ERBB2* and *EGFR* TPM expression levels for samples from normal brain and esophagus. To identify genes that were upregulated in only primary EAC and EAC brain metastasis groups, genes were removed in which the sum TPM expression levels for all samples was less than 300. From this list the top 500 upregulated genes were selected for primary EAC and the top 500 upregulated genes were selected from EAC brain metastasis. We used the David Gene Ontology website^66^ to find pathways upregulated in primary EAC and EAC brain metastasis.

### Hi-C Analysis

We performed Hi-C analysis using the 4D Nucleome Data (4DN) analysis pipeline (v0.2.7).^67^ Shortly, this pipeline performs pre-processing of data including reference-based alignment of reads to GRCh38 reference genome using bwa (v0.7.17). This pipeline also performs filtering and marking duplicates using pairtools (v0.2.2). Next the analysis pipeline employs Cooler (v0.8.3) which performs aggregation and normalization of data and produces cool files which were then utilized for downstream analysis. We utilized JuiceBox (v2.20.00)^68^ and set it in reconstruction mode to reconstruct the regions that spanned predicted ecDNA amplicons based on WGS results for Patient 7.

### Single-Cell Multi-ome Analysis

We leveraged both single-cell RNA sequencing (scRNA-seq) and single-cell ATAC sequencing (scATAC-seq) data from a single sample to infer copy number variations and tumor evolution. The 10x Cell Ranger pipeline was first implemented to process barcodes and perform single cell genome counting and Feature Barcode analysis to process the data. For scATAC-seq analysis, 10x Genomics Cell Ranger ARC (v2.0) facilitated the entire single-cell analysis pipeline, including filtering, transcript counting, cell clustering, and dimensionality reduction using UMAP. Initially, Seurat (v4/v5)^69^ was employed to generate a Seurat object from the Cell Ranger filtered feature matrix, where RNA transcripts were quantified, and cells were clustered based on the results of both scRNA-seq and scATAC-seq. Only cells that were common to both scATAC-seq and scRNA-seq datasets were retained, while others were excluded. Sc-type (v1.0)^70^ was utilized for the scRNA sequencing to perform cell clustering and automated cell type annotation. Unknown cell types were filtered out, and malignant cells were manually annotated. For scATAC-seq data, epiAneufinder (v0.1)^40^ was utilized to determine inferred copy numbers, allowing binning at 100 kb excluding blacklist cells (366 cells remained).

### Single-Nuclei Whole Genome Sequencing Analysis

Single-nuclei whole genome sequencing analysis was performed according to previous study^39^. Briefly, for single nuclei whole genome data, every well was aligned to the reference genome hg19 with Bowtie2.^71^ The output SAM files were then converted to BAM files and sorted using SAMtools. The sorted files were then converted back to SAM using SAMtools and Markdup was run to mark duplicates and then it was converted into varbin files utilizing runVarbins. The low-quality cells were removed. These low-quality cells are defined as cells that failed to pass the criteria “median bin count <10”. Normalized read counts per bin for each sample was produced to make the final heat map that shows the relative coverage per bin for each cell in each of our samples.^39^ We utilized ComplexHeatmap (v2.22.0)^72^ from R to produce the final heatmap figure which displayed the binned log2 ratio of coverage across the whole genome and across specific chromosomes. We used ASCAT.sc (v1)^73^ on BAM files for copy number calling. ASCAT.sc with adjusted ploidy was used to consider WGD results to plot the copy number per cell. We then plotted copy number per cell with *ERBB2* copy number on the x-axis and *EGFR* copy number on the y-axis utilizing a density scatter dot plot to show the copy number of these two genes in the same cell. We also performed pseudobulking of the single cell binned coverage ratio data by taking the average coverage for each bin to plot the pseudobulk coverage ratio and highlight the amplified regions with higher coverage.

### Xenium Single-Cell Spatial Transcriptomics Analysis

For spatial transcriptomics, we performed spatial analysis on FF and FFPE primary and brain metastasis tumor sections using the Xenium Analyzer (10x Genomics), our in-house machine. We followed the Xenium protocol as outlined by the manufacturer to ensure standardized processing and analysis. We analyzed adjacent primary and brain metastasis sections with two distinct gene panels. The first panel, the Xenium Human Brain Gene Expression Panel (v1, 10x Genomics), contained 266 genes, supplemented with a custom panel of 100 additional genes. The second panel, the Xenium Human Immuno-Oncology Profiling Panel (v1, 10x Genomics), included 380 genes. In total, we analyzed 34 tumor sections, comprising adjacent sections of five unique primary EAC tumors, eight unique EAC brain metastases, and two unique metastases from other sites.

Initial pre-processing of the data was performed by the Xenium Onboard Analysis software (10x Genomics) on the Xenium Analyzer. For further analysis of the data from the human brain panel we utilized Xenium Ranger (v1.7.0.2, 10x Genomics) to perform re-segmentation to modify the cell segments that were determined from the Xenium Onboard Analysis. The re-segmentation was set to 5 microns which is the standard size of a nucleus of a cell. We applied this re-segmentation process specifically to the brain panel Xenium data, which lacked cell segmentation markers due to the technology’s early stage of development. In contrast, the Xenium immune-oncology panel data included cell segmentation markers. While the analysis of these samples followed a similar approach to other samples, we did not perform cell re-segmentation, as the cells were already well-segmented. Instead, we applied additional filtering to remove clusters with a low mean transcript count, given the higher overall cell count. Further filtering ensured that only the highest-quality cells were retained for analysis.

We performed the remaining analysis using single-cell RNA-sequencing python-based packages using the same methods for all samples. We performed pre-processing of the data based on quality and number of transcripts per nucleus, utilizing AnnData (v0.10.7)^74^ structure and filtering using Scanpy (v1.9)^75^ python packages. We filtered cells out based on a minimum count threshold of 10 (40 for immune-oncology panel) which removes cells that have fewer transcripts counts than the threshold. Furthermore, we filtered genes out based on a minimum cell threshold of 5 (10 for immune-oncology panel) in which any genes that were in fewer than 5 cells were filtered out. We used the Scanpy package for normalization, adjusting transcript counts by scaling each cell’s total counts across all genes so that all cells have the same total count after normalization. We performed Scanpy dimensionality reduction using Leiden cell clustering to identify distinct cell clusters based on similarity of gene expression profiles. We visualized cell clusters using Scanpy UMAP (Uniform Manifold Approximation and Projection) to explore relationships between different cell clusters. We performed a Wilcoxon ranked statistical test of the genes in each cell cluster to determine the cell markers for each cluster. We then utilized the highest expressed genes in each cluster to manually annotate the cell clusters into unique cell types including immune cells based on the immune-oncology panel. We visualized the spatial plots using 10x Genomics Xenium Explorer (v3.1.1).

We utilized R to plot the log2+1 transcript counts for specific transcripts per sample and per cell type. We performed a statistical analysis using a Wilcoxon rank sum multiple hypothesis test with a p-value of 0.05 and a Benjamini-Hochberg adjustment to determine if the log2+1 transformed transcript count for specific genes differed between cell types or between different samples for the same cell type.

Additionally, to investigate the immune microenvironment we utilized the algorithm TLS-Finder (v1)^28^ to identify tertiary lymphoid structures and lymphoid aggregates, setting the detection radius to 160 microns. The algorithm defines a lymphoid aggregate as a structure of 50 or more T cells or B cells within the radius of detection, while a tertiary lymphoid structure is defined as having 50 or more T cells or B cells, along with the presence of dendritic cells.

To analyze ligand-receptor interactions for cytokines and chemokines within the Xenium immune panel datasets, we utilized CellChat (v2).^33^ This tool allowed us to assess cell-to-cell communication within our spatially resolved single-cell data.

