## Supplementary Figures for "Recurrent *ERBB2* alterations are associated with esophageal adenocarcinoma brain metastases"

### Supplemental Figures

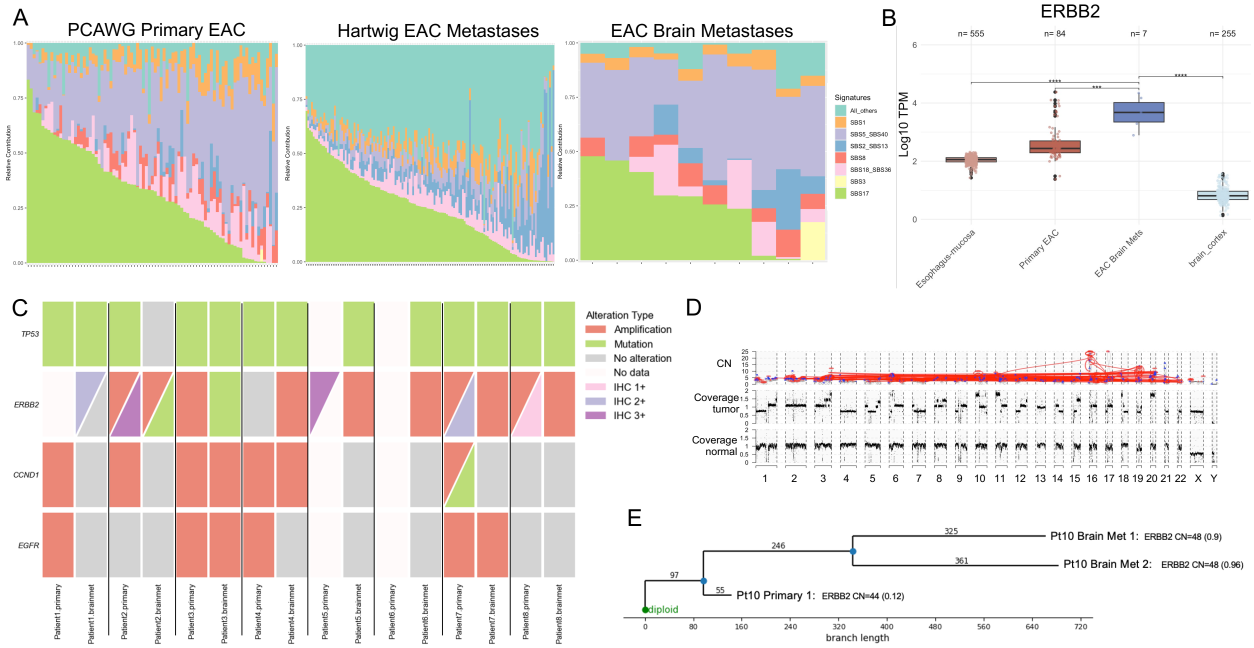

**Figure S1: *ERBB2* is a recurrent oncogene alteration in EAC brain metastases, Related to Figure 1**

(A) Relative contribution of COSMIC mutational signatures for PCAWG primary samples, Hartwig EAC metastases, and EAC brain metastases
(B) Box plot shows log10 TPM for *ERBB2* comparing TCGA primary EAC, EAC brain metastasis, and GTEX normal esophagus and brain tissues using a Wilcoxon rank sum test (p-value cutoff 0.05)
(C) Color boxes show alteration types indicated for primary and matched brain metastases samples for key driver genes including *TP53, ERBB2, CCND1*, and *EGFR.* HER2 IHC results is indicated for samples in which data was available
(D) A representative EAC brain metastasis (Patient 10) plot showing rearrangement, copy number (CN) and genome coverage
(E) Phylogenetic analysis of multi-region WGS data from a patient (Patient 10) showing early alterations in *ERBB2* gene. Locations of biopsies included with copy number for *ERBB2* and purity of tumor sample.

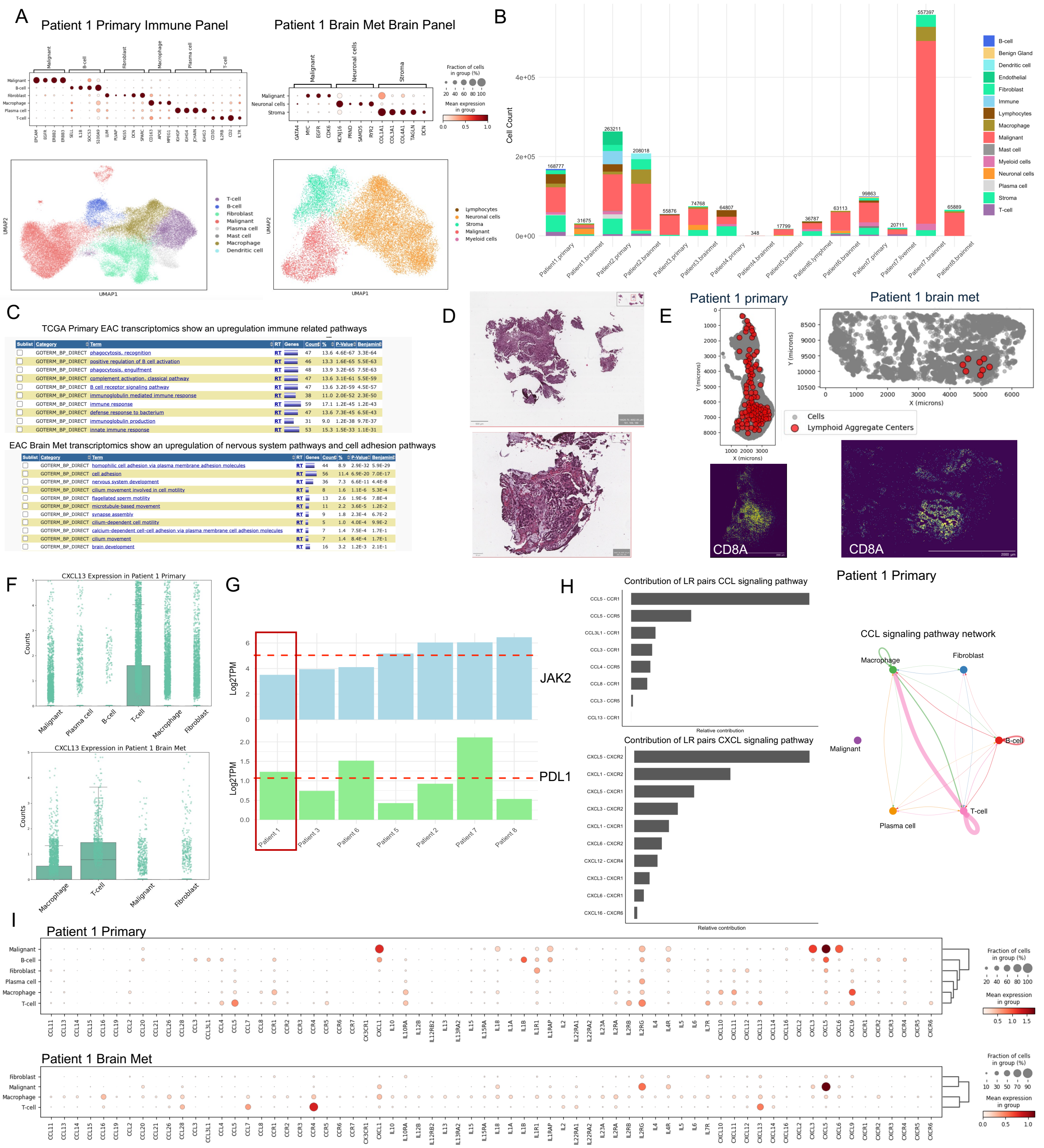
A
Patient 1 Primary Immune Panel
Patient 1 Brain Met Brain Panel
B
Cell Count
C
TCGA Primary EAC transcriptomics show an upregulation immune related pathways
EAC Brain Met transcriptomics show an upregulation of nervous system pathways and cell adhesion pathways
D
E
Patient 1 primary
Patient 1 brain met
Cells
Lymphoid Aggregate Centers
CD8A
CD8A
F
CXCL13 Expression in Patient 1 Primary
CXCL13 Expression in Patient 1 Brain Met
G
JAK2
PDL1
H
Contribution of LR pairs CCL signaling pathway
Contribution of LR pairs CXCL signaling pathway
Patient 1 Primary
CCL signaling pathway network
I
Patient 1 Primary
Patient 1 Brain Met

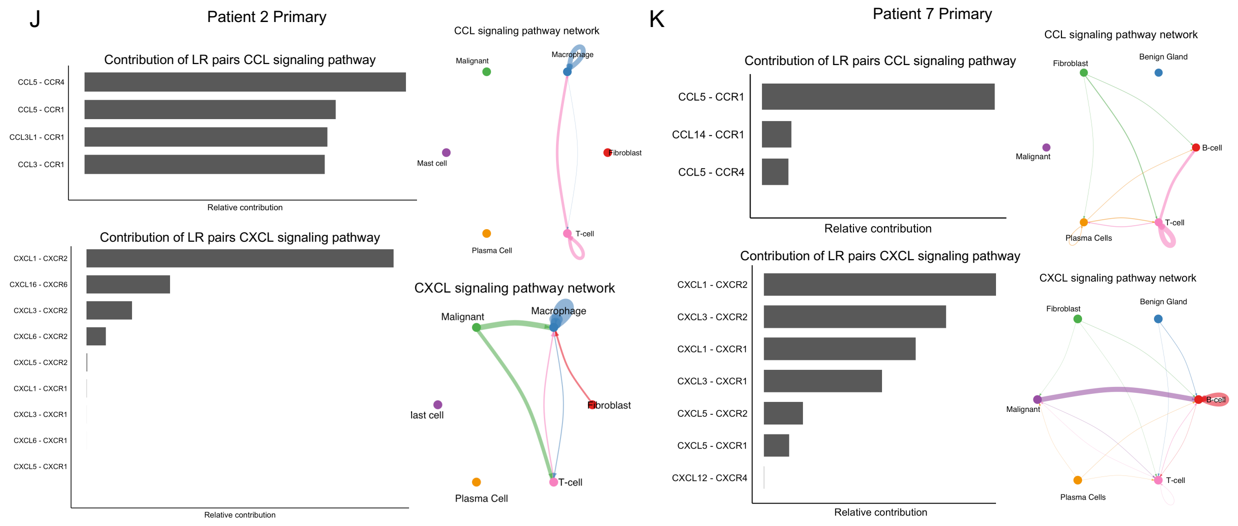


**Figure S2: Immune cell infiltration in primary tumors and high T cell infiltration in brain metastasis with *JAK2* deletion treated with immunotherapy, Related to Figure 2**

(A) (upper panel) Patient 1 primary immune panel and brain metastasis brain panel cell type marker gene dot plot and (lower panel) associated UMAPs
(B) Bar plot shows cell type count per patient with total cell count indicated above each bar and cell type indicated by different colors
(C) Results for gene ontology pathway enrichment analysis for TCGA Primary EAC cohort and EAC Brain Met cohort utilizing bulk RNA sequencing results
(D) Matched H&E images shown in Figure 2C from Patient 7 primary tumor display immune cell infiltration
(E) (upper panel) Patient 1 primary and brain metastasis spatial maps display the primitive tertiary lymphoid structures/lymphoid aggregates (LA) indicated by TLS-Finder algorithm and (lower panel) spatial density maps display *CD8A* expression
(F) Boxplot displays *CXCL13* expression from Xenium data in Patient 1 primary and brain metastasis samples separated by cell type
(G) Bar plot displays log2 TPM from bulk RNA sequencing data for *JAK2* and (*CD274*) PDL1 in our brain metastasis cohort. Bars are arranged from lowest to highest *JAK2* expression with the average expression for both genes shown as a dotted red line. Patient 1 brain metastasis is highlighted with red box.
(H) CellChat ligand receptor contribution analysis results for CCL (upper panel) and CXCL (lower panel) pathways for Patient 1 primary. (right) Circle plot displays the CCL signaling pathway network
(I) Cytokine expression marker gene dot plot by cell type shows the mean expression and fraction of cells expressing each cytokine by cell type for Patient 1 primary and brain metastasis samples
(J) CellChat ligand receptor contribution analysis results for CCL (upper panel) and CXCL (lower panel) pathways for Patient 2 primary. (right) Circle plot displays the CCL and CXCL signaling pathway network
(K) CellChat ligand receptor contribution analysis results for CCL (upper panel) and CXCL (lower panel) pathways for Patient 7 primary. (right) Circle plot displays the CCL and CXCL signaling pathway network

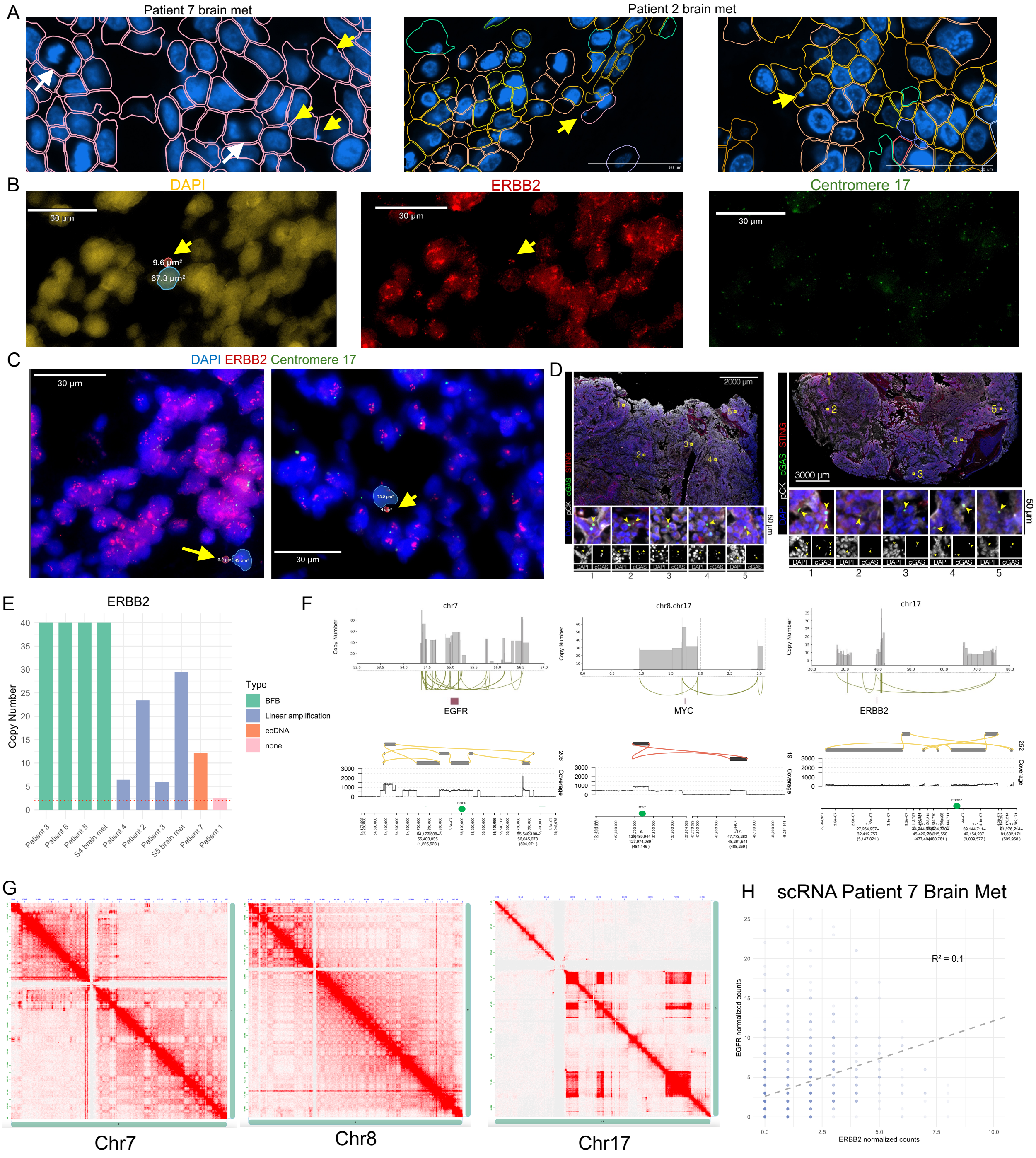


**Figure S3: Chromosomal instability of EAC brain metastasis tumors is marked by presence of micronuclei and ecDNA, Related to Figure 3**

(A) Example micronuclei (indicated by yellow arrows) and mitotic-like structures (indicated by white arrows) identified from Xenium in Patient 7 and Patient 2 brain metastasis samples showcasing the detection of genomic instability utilizing the DAPI staining of Xenium approach across tumor sections
(B) Fluorescence images of *ERBB2* FISH (red), centromere 17 (green), and DAPI (yellow) staining to detect *ERBB2* amplification and micronuclei in Patient 7 from matched images corresponding to Figure 3A. The sizes of micronuclei and adjacent nuclei are indicated

(C) Fluorescence images of *ERBB2* FISH (red), centromere 17 (green), and DAPI (blue) staining to detect *ERBB2* amplification and additional micronuclei examples on an adjacent section from Patient 7. The sizes of micronuclei and adjacent nuclei are indicated
(D) Immunofluorescence images show example micronuclei detected using cGAS and STING IF across various sections of the Patient 7 brain metastasis
(E) *ERBB2* copy number and alteration type for each patient determined using Amplicon Architect algorithm
(F) (top panel) Patient 7 ecDNA structures identified by Amplicon Architect (bottom panel) the same ecDNA species are detected via Decoil algorithm utilizing long-read sequencing
(G) Hi-C results from Patient 7 displays chromatin interaction patterns for chromosomes 7, 8, and 17 which shows high copy number levels resembling ecDNA amplifications
(H) Scatter plot with linear regression line and R-squared value displays *EGFR* and *ERBB2* co-expression from single cell RNA sequencing results

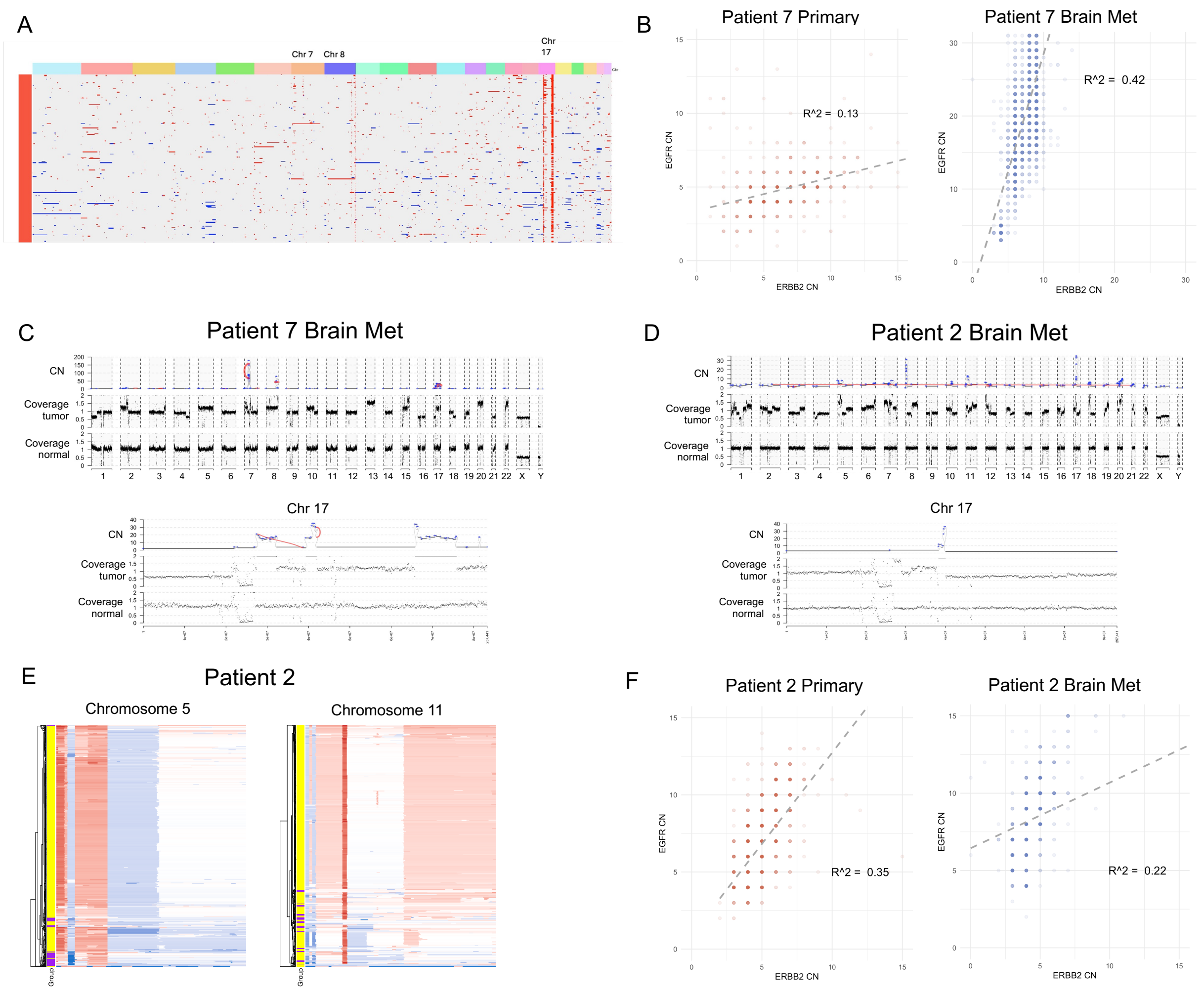


**Figure S4: Early and monoclonal metastatic seeding events, Related to Figure 4**

(A) scATAC heatmap displays inferred copy number gains in red and inferred copy number losses in blue in Patient 7 brain metastasis sample
(B) Scatter plot for with linear regression line displays *EGFR* and *ERBB2* copy number for each cell from single cell WGS results for Patient 7
(C) Patient 7 EAC brain metastasis plot showing rearrangement, copy number (CN) and genome coverage for whole genome and chromosome 17
(D) Patient 7 EAC brain metastasis plot showing rearrangement, copy number (CN) and genome coverage for whole genome and chromosome 17
(E) scWGS heatmap of log2 binned coverage ratio per chromosome for Patient 2 primary and brain metastasis samples.
(F) Scatter plot with linear regression line displays *EGFR* and *ERBB2* copy number for each cell from single cell WGS results for Patient 7
